# Assessment of SLC25A46 variants in Parkinson’s disease

**DOI:** 10.1101/2022.06.13.22276317

**Authors:** Hui Liu, Mohammad Dehestani, Mary B. Makarious, Sara Bandres-Ciga, Thomas Gasser, Jonggeol J. Kim

**Affiliations:** Department of Neurodegenerative Diseases, Hertie Institute for Clinical Brain Research, University of Tuebingen and German Center of Neurodegenerative Diseases (DZNE), Tuebingen 72076, Germany; Molecular Genetics Section, Laboratory of Neurogenetics, National Institute on Aging, National Institutes of Health, Bethesda, MD 20892, USA; Preventive Neurology Unit, Wolfson Institute of Preventive Medicine, Queen Mary University of London

**Keywords:** Parkinson’s disease, Genetics, risk factor, *SLC25A46*

## Abstract

The *SLC25A46* gene was recently reported to be associated with Parkinson’s disease (PD). Here, we comprehensively investigated the role of *SLC25A46* variants in PD patients of European ancestry and assessed susceptibility using whole-genome sequencing data from 1647 patients with PD and 1050 healthy controls. Burden analysis of rare non-synonymous variants across case-control individuals from whole-genome data did not find evidence of *SLC25A46* association with PD. Therefore, our results do not support a major role for *SLC25A46* in PD in the European population and large-scale sequencing studies of family trios are necessary to further evaluate the role of *SLC25A46* in PD etiology.

## Introduction

Many putative causal genes have been linked with Parkinson’s Disease (PD), and recently a possible link between compound heterozygous recessive *SLC25A46* variants and Parkinson’s disease (PD) have been suggested (Bitetto et al., 2020). Previous studies have shown that mutations in *SLC25A46*, encoding for a mitochondrial carrier protein, could cause peripheral neuropathy and optic atrophy (Charlesworth et al., 2016). Through whole-exome sequencing and gene screening, out of seven patients displaying parkinsonism accompanied with optic atrophy, two patients were found to have compound heterozygous *SLC25A46* variants (p.A401Sfs*17/ p.H137R and p.A176V/ p.K256R). The two carriers had parkinsonism symptoms onset at 43 years and 63 years respectively, and optic atrophy onset at 16 years and 52 years.

As part of the International Parkinson’s Disease Genomics Consortium’s (IPDGC) efforts to examine reported risk and causal factors for PD, we analyzed the role of *SLC25A46* variants in PD. We used publicly available whole-genome sequencing (WGS) data from the Accelerating Medicines Partnership - Parkinson’s disease initiative (AMP-PD) consisting of 1,647 PD patients (mean AAO 64.2 ± 9.6) and 1,050 neurologically healthy controls of European ancestry (mean age 60.3 ± 11.9) (www.amp-pd.org).

## Results

Our WGS analysis identified a total of 184 variants within the *SLC25A46* gene, 16 of which were coding variants, including 14 low-frequency or rare variants (MAF < 0.05). These coding variants included 7 synonymous and 9 non-synonymous variants (Table 1).

**Table 1.** Score test results of *SLC25A46 non-synonymous* coding variants in whole-genome sequencing data from PD and controls.

| Variant | rsID | Frequency (gnomAD) | Frequency (cases) | Frequency (controls) | OR | 95% CI | P-value |
| --- | --- | --- | --- | --- | --- | --- | --- |
| p.P50T | rs147648476 | 0.0003 | 0.001214 | 0 | 3.955 | 0.314 – 49.765 | 0.287 |
| p.E79K | rs200566665 | 0.0009 | 0.001821 | 0.002857 | 0.911 | 0.216 – 3.841 | 0.899 |
| p.T139N | rs202123515 | 0.0005 | 0.001214 | 0.000952 | 1.053 | 0.139 – 7.996 | 0.96 |
| p.V211M | rs114859074 | 0.0019 | 0.003947 | 0.002381 | 2.502 | 0.826 – 7.575 | 0.105 |
| p.R231Q | rs139039234 | - | 0 | 0.000476 | 0.349 | 3.70E-05 – 3284.354 | 0.821 |
| p.G249D | rs149585060 | 0.0002 | 0.000304 | 0 | 3.883 | 0.0439 – 343.029 | 0.553 |
| p.K256R | rs200725073 | 0.002 | 0.001821 | 0.002857 | 0.41 | 0.113 – 1.486 | 0.175 |
| p.L259R | rs141213807 | 0.0319 | 0 | 0.000476 | 0.31 | 0.00110 – 87.030 | 0.684 |
| p.E379D | rs150754737 | 0.0332 | 0.000304 | 0.000476 | 0.934 | 0.00414 – 211.095 | 0.98 |

CI, confidence interval; OR, odds ratio.

The variants p.E79K/p.V211M were present in compound heterozygosity in 1/1,050 controls and 1/1,647 cases (Fisher’s exact test, p = 1). No PD cases or controls were carriers of homozygous variants. Among the four coding variants reported by Bitetto et al, only the p.K256R variant was present in AMP-PD WGS data. Score tests were performed to assess the effect of individual variants on the risk of PD. Additionally, the cumulative effect of multiple rare *SLC25A46* variants on the risk of PD was assessed using gene-based burden analysis, including sequence kernel association tests (SKAT) and optimized SKAT (SKAT-O) using the RVTESTS package v.2.1.0 (Zhan et al., 2016). Although we assume the limitation that our analyses might be underpowered, we did not detect enrichment of rare variants in PD cases versus controls (Table 2) and the score test did not show significant differences in the frequency of *SLC25A46* variants between PD cases and controls (Table 1).

**Table 2.**
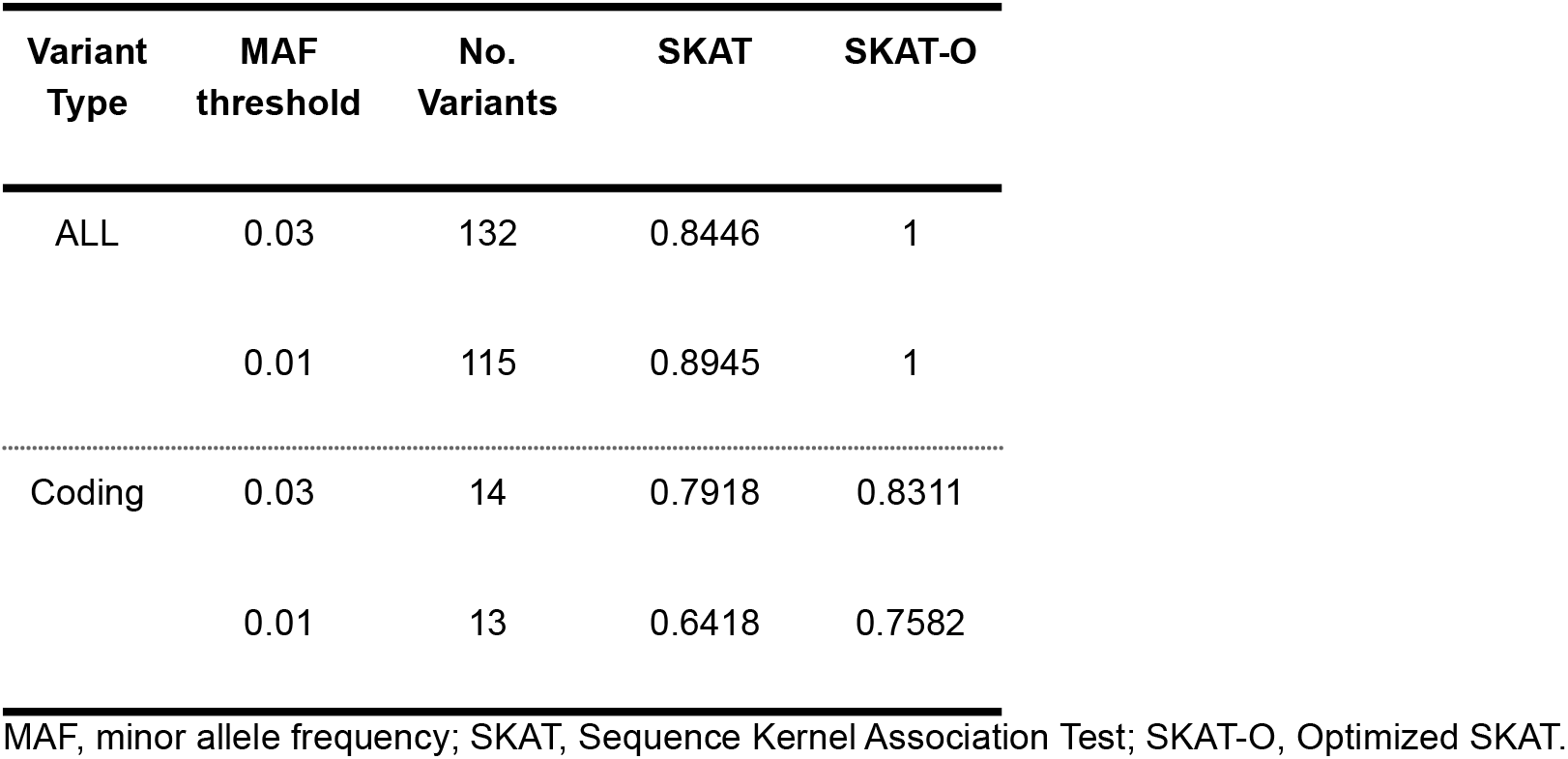
*SLC25A46* burden analyses using whole-genome sequencing data

## Conclusion

In summary, we did not find any enrichment of *SLC25A46* compound heterozygous or homozygous variants in PD cases compared to controls. There was no significant difference in the frequency of the p.K256R, reported variant, in PD cases and controls. The frequencies observed in cases are very similar to those observed in the general population, as reported in gnomAD (https://gnomad.broadinstitute.org/). Based on our analysis, no significant burden of *SLC25A46* rare variants was found to increase PD risk. The previously identified mutations are linked to a very unusual phenotype that presents with parkinsonism and optic atrophy, but both phenotypes may not be etiologically related to PD. However with only 1,647 PD patients 1,050 controls, our results may be too underpowered to detect an association between these variants and PD. Both this study and previous study focused exclusively on the European population, and it may be helpful to explore the potential relationship between this gene and PD in a more diverse context. In conclusion, our findings do not suggest that *SLC25A46* is a causal gene or risk factor for PD, but to minimize potential type-II error large-scale sequencing studies of family trios are necessary to further evaluate the role of *SLC25A46* in PD etiology.

## Methods

The publicly available AMP-PD data was used for this analysis. Cohort, sample characteristics, and quality control pipeline for AMP-PD data are described in https://amp-pd.org/whole-genome-data. Detailed description of the analysis pipeline is provided as supplementary information and also available in the Github repository here: https://github.com/ipdgc/IPDGC-Trainees/blob/master/SLC25A46.md. In short, PLINK 1.9 was used to isolate the region of interest, PLINK 2 was used to extract allele frequency and count, and ANNOVAR (version 2018Apr16) was used to annotate the variants. We used RVTESTS package v.2.1.0 to perform score test and gene-based burden analyses. Both score test and gene-based burden analysis were adjusted by sex, age, education level, 10 principal components (PCs) to account for population stratification, and cohort. Significance of the compound heterozygosity variant was determined using the Fisher test, which was performed in R version 3.6.

## Data Availability

All data produced in the present study are available upon reasonable request to the authors

## Acknowledgments

We would like to thank all of the subjects who donated their time and biological samples to be part of this study. We would also like to thank all members of the International Parkinson Disease Genomics Consortium (IPDGC). For a complete overview of members, acknowledgments, and funding, please see http://pdgenetics.org/partners. We would like to thank the Accelerating Medicines Partnership initiative (AMP-PD) for the publicly available whole-genome sequencing data release version 1.0, November 2019, including cohorts from the BioFIND study, the NINDS Parkinson’s disease Biomarkers Program (PDBP), and MJFF Parkinson’s Progression Marker Initiative (PPMI). Clinical data and biosamples used in preparation of this article were obtained from the Fox Investigation for New Discovery of Biomarkers (BioFIND), the Harvard Biomarker Study (HBS), the Parkinson’s Progression Markers Initiative (PPMI), and the Parkinson’s Disease Biomarkers Program (PDBP). BioFIND is sponsored by The Michael J. Fox Foundation for Parkinson’s Research (MJFF) with support from the National Institute for Neurological Disorders and Stroke (NINDS). The BioFIND Investigators have not participated in reviewing the data analysis or content of the manuscript. For up-to-date information on the study, visit www.michaeljfox.org/biospecimens. The Harvard Biomarkers Study (HBS) is a collaboration of HBS investigators [full list of HBS investigators found at https://www.bwhparkinsoncenter.org/biobank] and funded through philanthropy and NIH and Non-NIH funding sources. The HBS Investigators have not participated in reviewing the data analysis or content of the manuscript. PPMI – a public-private partnership – is funded by the Michael J. Fox Foundation for Parkinson’s Research and funding partners. Other funding partners include a consortium of industry players, non-profit organizations and private individuals, and include the following (in alphabetical order): Abbvie, Allergen, Amathus Therapeutics, Avid Radiopharmaceuticals, Biogen, BioLegend, Bristol-Myers Squibb, Celgene, Denali, GE Healthcare, Genentech, GlaxoSmithKline, Golub Capital, Handl Therapeutics, Insitro, Janssen Neuroscience, Eli Lilly, Lundbeck, Merck, Meso Scale Discovery, Pfizer, Piramal, Prevail Therapeutics, Roche, Sanofi Genzyme, Servier, Takeda, Teva, UCB, Verily, and Voyager Therapeutics. Industry partners are contributing to PPMI through financial and in-kind donations and are playing a lead role in providing feedback on study parameters through the Partner Scientific Advisory Board (PSAB). Through close interaction with the study, the PSAB is positioned to inform the selection and review of potential progression markers that could be used in clinical testing. The PPMI Investigators have not participated in reviewing the data analysis or content of the manuscript. For up-to-date information on the study, visit www.ppmi-info.org. Parkinson’s Disease Biomarker Program (PDBP) consortium is supported by the National Institute of Neurological Disorders and Stroke (NINDS) at the National Institutes of Health. A full list of PDBP investigators can be found at https://pdbp.ninds.nih.gov/policy. The PDBP Investigators have not participated in reviewing the data analysis or content of the manuscript. As registered users of the AMP-PD, HL, MBM, SJL, and JJK have access to individual-level data. This work was supported in part by the Intramural Research Programs of the National Institute of Neurological Disorders and Stroke (NINDS), the National Institute on Aging (NIA), and the National Institute of Environmental Health Sciences both part of the National Institutes of Health, Department of Health and Human Services; project numbers 1ZIA-NS003154, Z01-AG000949-02 and Z01-ES101986. In addition, this work was supported by the Department of Defense (award W81XWH-09-2-0128), and The Michael J Fox Foundation for Parkinson’s Research.

## Conflicts of Interest

The authors declare that they have no conflict of interest.

## Notes

### Competing Interest Statement

The authors have declared no competing interest.

### Funding Statement

This study did not receive any funding

### Author Declarations

The study used ONLY openly available human data that were originally located at AMP-PD(https://amp-pd.org/).

